## Supplementary material for "Clinical outcomes associated with schistosome infection and alcohol use: a systematic scoping review"

**Number of hits from each database searched**

| PubMed/MEDLINE | | |
| --- | --- | --- |
| Search number | Search term | Number of results |
| 1 | (Schistosom* OR Bilharzia OR snail* fever) | 39,520 |
| 2 | (alcohol OR drink*) | 1,248,528 |
| 3 | 1 AND 2 | 614 |

| Embase (via Ovid) | | |
| --- | --- | --- |
| Search number | Search term | Number of results |
| 1 | (Schistosom* OR Bilharzia OR snail* fever).mp. | 41,620 |
| 2 | (alcohol OR drink*).mp. | 923,069 |
| 3 | 1 AND 2 | 735 |

[mp = title, abstract, heading word, drug trade name, original title, device manufacturer, drug manufacturer, device trade name, keyword heading word, floating subheading word, candidate term word]

| Global Health (via Ovid) | | |
| --- | --- | --- |
| Search number | Search term | Number of results |
| 1 | (Schistosom* OR Bilharzia OR snail* fever).mp. | 39,224 |
| 2 | (alcohol OR drink*).mp. | 249,939 |
| 3 | 1 AND 2 | 286 |

| Global Index Medicus | | |
| --- | --- | --- |
| Search number | Search term | Number of results |
| 1 | (Schistosom* OR Bilharzia OR “snail fever”) | 49,490 |
| 2 | (alcohol OR drink*) | 40,346 |
| 3 | 1 AND 2 | 417 |

| Web of Science | | |
| --- | --- | --- |
| Search number | Search term | Number of results |
| 1 | (Schistosom* OR Bilharzia OR “snail fever”) | 39,496 |
| 2 | (alcohol OR drink*) | 1,009,328 |
| 3 | 1 AND 2 | 306 |
